# Genetic correlations between COVID-19 and a variety of traits and diseases

**DOI:** 10.1101/2020.12.18.20248319

**Authors:** Xiao Chang, Yun Li, Kenny Nguyen, Huiqi Qu, Yichuan Liu, Joseph Glessner, Patrick M A Sleiman, Hakon Hakonarson

## Abstract

We analyzed GWAS results released by COVID-19 Host Genetics Initiative, UK biobank and GWAS Catalog to explore the genetic overlap between COVID-19 and a broad spectrum of traits and diseases. We validate previously reported medical conditions and risk factors based on epidemiological studies, including but not limited to hypertension, type 2 diabetes and obesity. We also report novel traits associated with COVID-19, which have not been previously reported from epidemiological data, such as opioid use and educational attainment. Taken together, this study extends our understanding of the genetic basis of COVID-19, and provides target traits for further epidemiological studies.

## Introduction

The ongoing coronavirus disease 2019 (COVID-19) break has posed an extraordinary threat to global public health. Patients with certain underlying medical conditions such as obesity, hypertension, and diabetes are at increased risk for poor outcome in COVID-19 ^1^. Given high genetic heritability of the aforementioned conditions, their shared genetic factors may play a crucial role in the severity of COVID-19. Indeed, a recent genome-wide association study (GWAS) of COVID-19 has reported two genomic loci associated with severe COVID-19, indicating a strong genetic influence on the severity of COVID-19 ^2^. Here, we analyzed GWAS results released by COVID-19 Host Genetics Initiative ^3^, UK biobank and GWAS Catalog to explore the genetic overlap between COVID-19 and a broad spectrum of traits and diseases.

## Methods

Summary statistics of selected COVID-19 GWAS (sample size > 30,000 and percentage of Europeans > 90%) were downloaded from COVID-19 Host Genetics Initiative including A2_ALL (very severe respiratory confirmed COVID-19 against population), B2_ALL (hospitalized COVID-19 against population), B2_ALL_eur (hospitalized COVID-19 against population in Europeans), C2_ALL_eur (COVID-19 against population in Europeans). GWAS summary statistics of selected diseases/traits were downloaded from UK biobank and GWAS Catalog ^4^. Genetic correlation r_g_ between COVID-19 and interested diseases/traits were estimated by LD score regression (LDSC) using GWAS summary statistics that overlap with HapMap3 SNPs as recommended ^5^. Pre-computed linkage disequilibrium scores for HapMap3 SNPs calculated based on European-ancestry individuals from the 1000 Genomes Project were used in the analysis.

## Results

We first investigated genetic correlations between COVID-19 and 1555 diseases/traits from the analysis of UK biobank data by Neale lab (http://www.nealelab.is/uk-biobank/). Our results are consistent with the epidemiological observation that BMI is significantly associated with severe or hospitalized COVID-19 (A2_ALL, rg = 0.24, *P* = 3.35 × 10^−6^; B2_ALL, rg = 0.39, *P* = 3.33 × 10^−7^). COPD (chronic obstructive pulmonary disease), heart diseases, hypertension, diabetes and smoking status exhibit substantial magnitude of genetic correlation with COVID-19, though statistical significance do not pass the strict threshold after adjustment for multiple testing (Table 1). Collectively, diseases of the circulatory system, diseases of the digestive system, and diseases of the musculoskeletal system and connective tissue are significantly associated with severe or hospitalized COVID-19 (Table 1). In agreement with this, a number of medication-taking traits linked to obesity, diabetes, hypertension and digestion display modest correlation with COVID-19 (Table 1). Tramadol, an opioid pain medication, is significantly correlated with hospitalized COVID-19 (B2_ALL, rg = 0.65, *P* = 1.26 × 10^−5^). Interestingly, our results indicated a significant negative correlation between severe or hospitalized COVID-19 and educational attainment related traits including college or university degree (A2_ALL, rg = -0.24, *P* = 8.51 × 10^−7^; B2_ALL, rg = -0.32, *P* = 1.78 × 10^−6^) and fluid intelligence score (A2_ALL, rg = -0.25, *P* = 2.40 × 10^−5^; B2_ALL, rg = -0.26, *P* = 7.66 × 10^−5^). Hospitalized COVID-19 (B2_ALL, rg = 0.73, *P* = 8.60 × 10^−6^) is also significantly correlated with panic attacks.

**Table 1.**
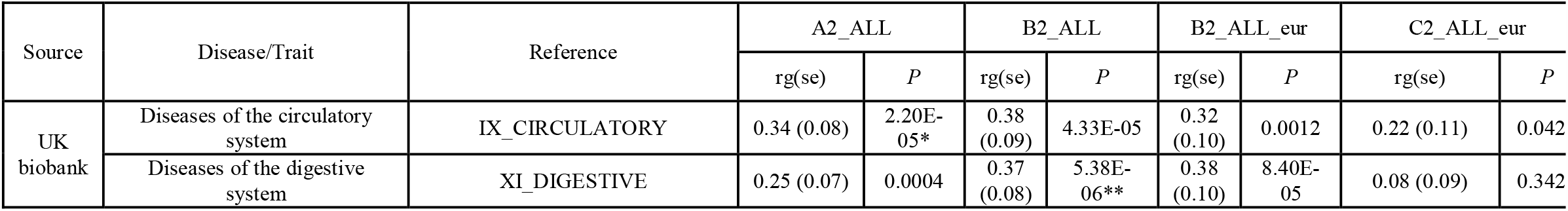
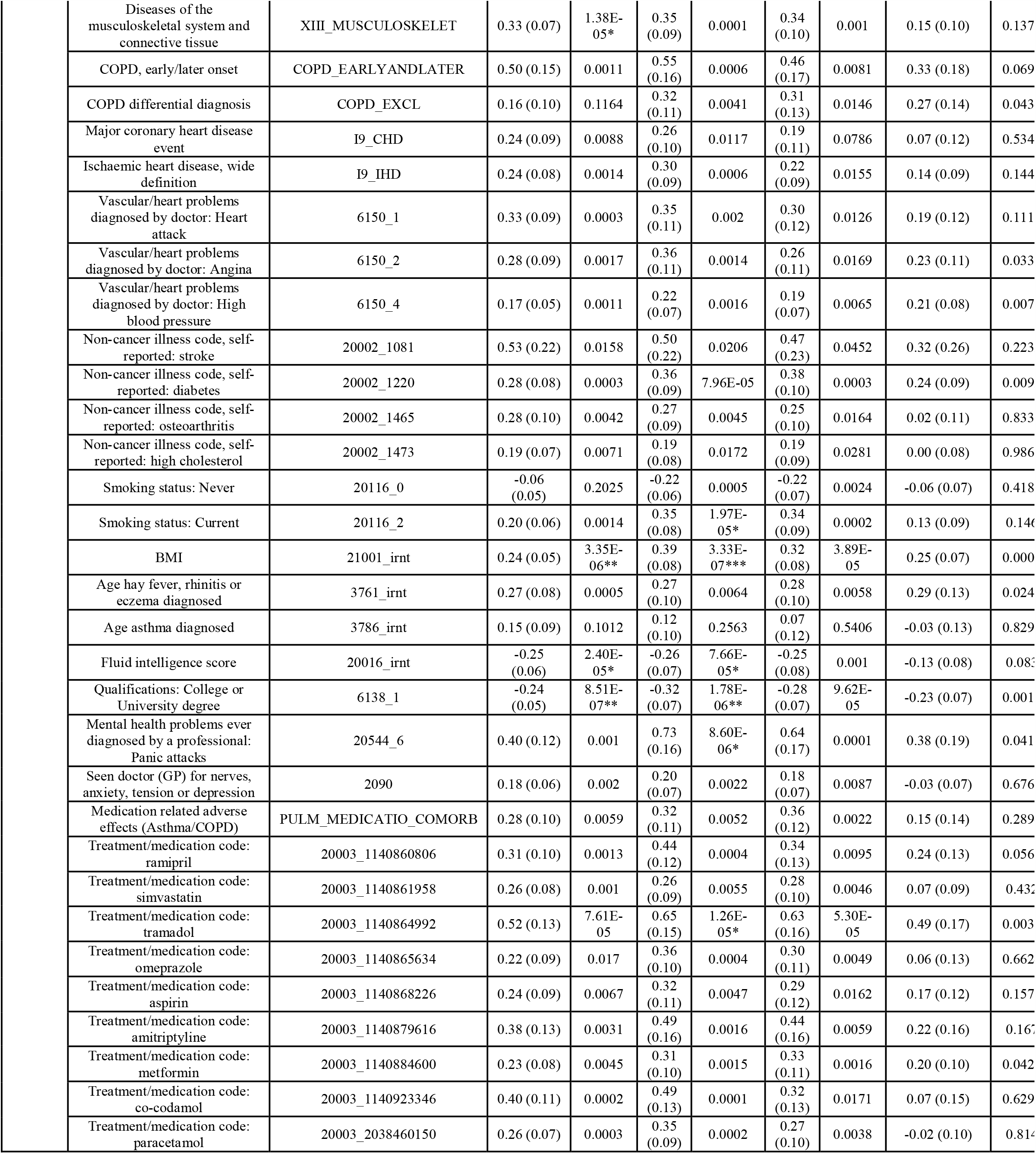
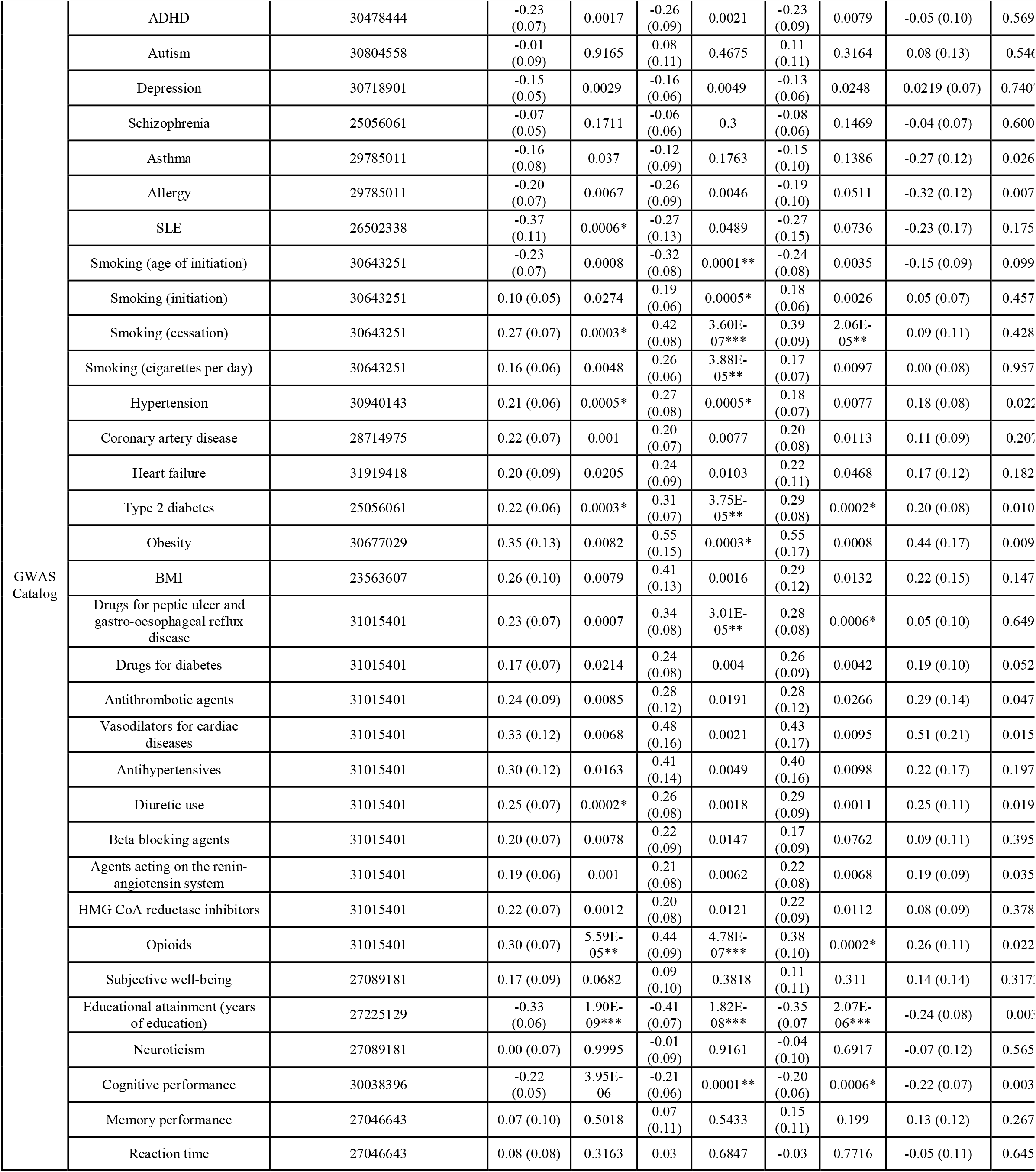
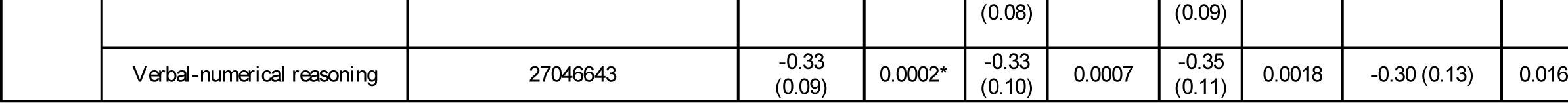
Genetic correlations between COVID-19 and a variety of diseases and other medically relevant traits

We next estimated genetic correlations between COVID-19 and 80 diseases/traits from GWAS Catalog ^4^. Consistently, hypertension, type 2 diabetes and obesity are significantly associated with severe or hospitalized COVID-19 (Table 2). Coronary artery disease, heart failure and BMI are also modestly associated with severe COVID-19. Likewise, medication-taking traits related to obesity, diabetes, hypertension and digestion are modestly associated with COVID-19 such as drugs for diabetes, and antihypertensives (Table 2). Significant correlations are also found between hospitalized COVID-19 and drugs for peptic ulcer and gastro-oesophageal reflux disease (B2_ALL, rg = 0.34, *P* = 3.01 × 10^−5^; B2_ALL_eur, rg = 0.28, *P* = 0.0006), diuretic use and very severe respiratory confirmed COVID-19 (A2_ALL, rg = 0.25, *P* = 0.0002), opioids and severe or hospitalized COVID-19 (A2_ALL, rg = 0.30, *P* = 5.59 × 10^−5^; B2_ALL, rg = 0.44, *P* = 4.78 × 10^−7^; B2_ALL_eur, rg = 0.38, *P* = 0.0002). In consistent with the findings from UK biobank data, a strong negative genetic correlation between educational attainment and severe or hospitalized COVID-19 (A2_ALL, rg = -0.33, *P* = 1.90 × 10^−9^; B2_ALL, rg = -0.41, *P* = 1.82 × 10^−8^; B2_ALL_eur, rg = -0.35, *P* = 2.07 × 10^−6^) is observed. Further analyses of brain function and personality traits show COVID-19 is significantly correlated with cognitive performance and verbal-numerical reasoning, but not memory performance, reaction time or neuroticism (Table 1). In addition, very severe respiratory confirmed COVID-19 (A2_ALL, rg = -0.37, *P* = 0.0006) is significantly correlated with systemic lupus erythematosus.

## Discussion

Our genetic correlation results between COVID-19 and a variety of traits and diseases confirm medical conditions and risk factors reported from epidemiological studies such as hypertension, type 2 diabetes and obesity. However, as far as we are aware, the association between opioids and COVID-19 has not previously been reported using epidemiological data. As side effects associated with chronic opioid use at high doses may affect the immune system and increase the risk of pneumonia, there is an urgent need to evaluate the relationship between COVID-19 severity and opioid use by epidemiological studies ^6^. Also, patients using chronic opioids may be considered a vulnerable group for careful monitoring. In addition, our results suggest that immune pathways involved in systemic lupus erythematosus may also play an important role in the severity of COVID-19. Finally, the observed negative correlation between COVID-19 and educational attainment reflects an indirect link mediated by environment or human behavior. For example, patients with different educational levels may differ in diet choices (BMI) or smoking status. This study provides novel information on underlying conditions that might increase the risk of severe illness of COVID-19. Added epidemiological studies are warranted to further evaluate these findings.

Genetic correlation estimates, standard errors and p-values for COVID-19 and selected diseases/traits. The “Reference” column lists the phenotype code for the GWAS from UK biobank, and PMID (PubMed ID) for the GWAS from GWAS Catalog. The *P*-values are uncorrected *P*-values. Bonferroni correction was used for correction of multiple testing by the number of tests (1555 for the analysis of data from UK biobank, and 80 for the analysis of data GWAS Catalog). *** adjusted *P*-value□≤□0.001, ** adjusted *P*-value□≤□0.01, * adjusted *P*-value□≤□0.05. BMI = body mass index; SLE = systemic lupus erythematosus; rg = genetic correlation estimates; se = standard error, A2_ALL = very severe respiratory confirmed COVID-19 vs population, B2_ALL = hospitalized COVID-19 vs population, B2_ALL_eur = hospitalized COVID-19 vs population (European samples only), C2_ALL_eur = COVID-19 vs population (European samples only).

## Data Availability

Data analyzed in this paper were obtained from public databases.

